# COVID-19 Vaccine Hesitancy among the Adult Population in Bangladesh: A Nationally Representative Cross-sectional Survey

**DOI:** 10.1101/2021.04.23.21255844

**Authors:** Mohammad Bellal Hossain, Md. Zakiul Alam, Md. Syful Islam, Shafayat Sultan, Md. Mahir Faysal, Sharmin Rima, Md. Anwer Hossain, Abdullah Al Mamun

**Author notes:** Corresponding Author: **Mohammad Bellal Hossain, PhD**.

## Abstract

**Introduction:** The study related to the COVID-19 vaccine hesitancy is scanty in the context of Bangladesh, despite the growing necessity of understanding the mass people’s vaccination-related behavior. Thus, the present study was conducted to assess the prevalence of the COVID-19 vaccine hesitancy and its associated factors in Bangladesh to fill the knowledge gap.

**Methodology:** This study adopted a cross-sectional study design to collect data from 1497 respondents using online (Google forms) and face-to-face interviews. We employed descriptive statistics and multiple hierarchical linear regression analysis.

**Findings:** The prevalence of vaccine hesitancy was 41.1%. Men had less hesitancy (β = -0.046, p = 0.030) than women. The Muslims (β = 0.057, p = 0.009) and the respondents living in the city corporation areas (β = 0.132, p <0.001) had more hesitancy. There was significant variation in vaccine hesitancy by administrative divisions (geographic regions). The vaccine hesitancy tended to decrease with increasing knowledge about the vaccine (β = -0.072, p=0.001) and the vaccination process (β= -0.058, p = 0.018). On the other hand, hesitancy increased with the increased negative attitudes towards vaccine (β = 0.291, p <0.001) and conspiracy beliefs towards the COVID-19 vaccine (β = 0.105, p=0.004). The perceived severity of the COVID-19 (β = -0.079, p=0.002) and perceived benefits of COVID-19 vaccination (β = -0.180, p=0.001) were negatively associated with hesitancy, while perceived barriers (β = 0.180, p <0.001) were positively associated. The participants were more hesitant to accept the vaccine from a specific manufacturer.

**Conclusion:** This study emphasizes that negative attitudes and conspiracies towards the COVID-19 vaccine should be reduced through effective communications and contracting with additional vaccine manufacturers should be prioritized. The barriers like online registration for receiving the COVID-19 vaccination need to be removed, and initiatives like text message service using the mobile phone operator can be introduced.

**Highlights:**

- About 41% of the respondents had had hesitancy to accept the COVID-19 vaccine.
- The hesitancy increased with negative attitudes about vaccines and conspiracy beliefs.
- Perceived barriers to receive the vaccine were increasing vaccine hesitancy.
- Perceived severity of the COVID-19 decreased the vaccine hesitancy.
- Perceived benefits of receiving the COVID-19 vaccine decreased the vaccine hesitancy.

## 1. Introduction

Vaccination is one of the most significant progress in the public health era though anti-vaccination attitudes, behavior, and associated misconceptions are prevalent worldwide [1]. The evidence shows that the effectiveness of vaccination programs has been affected by *vaccine hesitancy* [2], where hesitancy has been defined as “delay in acceptance or refusal of vaccination despite the availability of vaccination services” [3]. Hesitancy regarding the Coronavirus diseases 19 (COVID-19) vaccination is prominently visible around the world [4] in such a period when the effort towards reaching herd immunity has been targeted to achieve through the mass vaccination coverage [5].

The Government of Bangladesh (GoB) has launched the biggest-ever mass vaccination program nationwide to vaccinate 80% (over 130 million) of the country’s total population with the COVID-19 vaccines in four stages [6] though nearly 34% population are below 18 years old [7]. The GoB has published a national deployment and vaccination plan for the COVID-19 vaccination that requires an online registration to receive the COVID-19 vaccine. According to the plan, the GoB has targeted to vaccinate 39.5 million population aged 40 years and above and frontline workers in the first phase. However, as of 17 April 2021, only 18% of the targeted people have been registered so far to receive vaccination, of whom 96% received the vaccine with a significant variation by gender (male 63%, female 37%) and administrative regions (Dhaka (26%) received more vaccination than other administrative divisions) [8]. Simultaneously, incidents about the lack of interest among the mass people about the vaccine uptake and lack of response about the registration process have been repeatedly reported in the media, showing vaccine hesitancy among the mass population.

The studies conducted to explore the COVID-19 vaccine hesitancy has shown that various socio-economic and demographic variables, different constructs of health belief model (HBM) [9,10], level of knowledge related to vaccine and vaccination process [11,12], attitude towards COVID-19 vaccination [12–14], conspiracy beliefs regarding the origin, effectiveness, and consequences of receiving vaccines [15–17], preventive behavioral practices related to COVID-19 [18,19], newness, safety, and probable side effects of the vaccine [20,21] have been primarily responsible for vaccine hesitancy around the world.

The study related to vaccine hesitancy, be it is COVID-19 or any other diseases, is scanty in the context of Bangladesh, despite the growing necessity of understanding the mass people’s vaccination-related behavior. Only one study has so far been conducted to assess the COVID-19 vaccine hesitancy, which has reported a 32.5% prevalence of vaccine hesitancy [14]. Thus, the present study was conducted to assess the prevalence of the COVID-19 vaccine hesitancy and its associated factors in Bangladesh to fill the knowledge gap.

## 2. Materials and Methods

### 2.1 Study Design and Data Collection

This study adopted a cross-sectional research design. We used the following formula to calculate the sample size:(Z^2^ pq/e^2^)Deff*NR. We used Z-score for 95% confidence interval (Z=1.96), prevalence (p) of willingness to accept a COVID-19 vaccine from an earlier study (p = 0.325) [14], margin of error (e = 0.03); design effect (Deff=1.6) for sampling variation; and a non-response rate (NR=10%). The calculated sample size was 1635. However, during the interview, 112 respondents did not consent to take part in the study. The response rate was 93.1 percent. We also had to exclude 26 respondents who responded that they did not know about the COVID-19 vaccine. Thus, the final sample for this study was 1497 for analysis. The data is now placed in the Mendeley open research data repository [22].

Data were collected between 1-7 February 2021. We chose this time for collecting the data as the GoB received two million doses of COVISHIELD (AstraZeneca) vaccine on 21 January 2021 as a gift from the Government of India and another 5 million from the Serum Institute of India on 25 January 2021 as a partial consignment of the 30 million doses of procured vaccination. On 27 and 28 January, the GoB vaccinated 567 frontline health workers and scheduled 7 February to start mass vaccination following the guideline.

Data were collected from all the eight administrative divisions of Bangladesh to make the study nationally representative. The samples were proportionately distributed to the population size of the divisions. We collected data using both online and face-to-face interviews. One-third of the respondents took part in this study online. The online data were collected through Google Forms using the Bengali language. The participants to whom the survey link was sent through e-mail, WhatsApp, or Facebook were requested to fill-up the form and circulate the link in their network to reach more people. The research team members circulated the survey link in their respective professional and social networks through the snowball process. The online link was valid for three days. The online data were downloaded, and divisional distribution was assessed. Data were then collected from the remaining sample size for each division through a face-to-face interview. Four days were allocated to collect data using a face-to-face interview. We selected two districts randomly from each of the divisions for collecting data through a face-to-face interview. Within each district, the samples were distributed proportionately according to the rural-urban distribution. Population aged 18 years and above, living in Bangladesh, and know about the COVID-19 vaccine were the selection criteria for the face-to-face interview. However, for the online survey, the ability to read and write and internet use were added with the face-to-face interview selection criteria.

### 2.2. Measures

#### Outcome Variable: Vaccine Hesitancy

We used two questions to measure this study’s outcome variable, which is the COVID-19 vaccine hesitancy. We asked the respondents what they will do if they get the chance of taking the COVID 19 vaccine for free? The responses to this question were: 1= Surely, I will take it; 2= Probably I will take it; 3= I will delay taking it; 4= I am not sure what I will do; 5= Probably I will not take it; 6= Surely, I will not take it. The second question was what they would do if their family or friends think of taking COVID 19 vaccine? The responses to this question were: 1= Strongly encourage them; 2=Encourage them; 3=Ask them to delay getting the vaccine; 4=I will not say anything about it; 5=Discourage them to take vaccine; 6=Forbid them to take the vaccine. The Cronbach Alpha (α) of these two items was 0.833, which shows good internal consistency. We combined these two items and made the level of vaccine hesitancy (ranges 2 to 12), where a higher score indicates higher hesitancy toward the COVID-19 vaccine.

#### Independent Variables

##### Socio-economic and Demographic Variables

We included the following socio-economic and demographic variables as the independent variables of this study: age, sex, religion, marital status, educational attainment, place of residence, administrative division, occupation, number of household members, household income.

##### Behavioral Practice to Prevent COVID-19

We measured preventive behavioral practices related to COVID-19 using three items. The total score of these items ranged between 1 and 12, with a higher score indicating higher better preventive practices with the Cronbach alpha (α=0.857).

##### Knowledge about the COVID-19 Vaccine

We assessed the knowledge related to the COVID-19 vaccine using four questions. The total score of these items ranged between 1 and 20, with a higher score indicating higher knowledge with the Cronbach alpha (α=0.643).

##### Knowledge about the Vaccination Process

Knowledge about the COVID-19 vaccination process was measured by six binary (yes=1, no=0) questions. The Cronbach alpha (α) of these six questions was 0.765, which shows good internal consistency. The higher the scores, suggest better knowledge.

##### COVID-19 Vaccine Conspiracy

Conspiracy related to the COVID-19 vaccine was measured using nine Likert scale items (α=0.716). The total score of these items ranged between 9 and 45, where a higher score shows higher conspiracy toward the COVID-19 vaccine.

##### Attitude towards COVID-19 Vaccine

COVID-19 vaccine-related attitudes (α=0.739) was assessed using six Likert-type items. The total score of attitudes toward the COVID-19 vaccine ranged between 6 and 30, where a higher score of this scale indicates higher negative attitudes toward the COVID-19 vaccine.

##### Health Belief Model

The classical health belief model (HBM) consists of the following components: perceived susceptibility, perceived severity, perceived benefits, and perceived barriers.

##### Perceived Susceptibility

Two five-point Likert scale questions were used to measure the perceived susceptibility (α=0.657) of the COVID-19.

##### Perceived Severity

Perceived severity of the COVID-19 was measured using two Likert scale questions, which had an α of 0.612.

##### Perceived Benefits

Perceived benefits (α=0.841) of the COVID-19 vaccination were measured using three five-point Likert scale questions. Perceived Barriers: The perceived barriers (α=0.700) of getting the COVID-19 vaccination were measured using six five-point Likert scale questions.

#### 2.3. Statistical Analysis

We first employed univariate descriptive statistical analysis [percentage, mean, and standard deviation (SD)]. We also applied the accuracy test of each scale, where we divided the mean score of a particular scale by the total score of that scale. The independent sample t-test, one-way ANOVA, and Pearson’s product-moment correlation were used to estimate the bivariate level statistics. The statistically significant (*p* ≤ 0.05) variables of the bivariate level were entered into the hierarchical multiple linear regression model after checking the assumptions and multicollinearity. We analyzed the data using the Statistical Product and Service Solutions (SPSS) software, version 26.

### 2.4. Ethical Approval

We took ethical approval from the National Research Ethics Committee of the Bangladesh Medical Research Council (BMRC). Participation in this study was completely voluntary, and no incentive was provided to the participants. The respondents were informed about the potential scopes and implications of the findings of this study and were requested to participate voluntarily.

## 3. Results

### 3.1. Characteristics of the Participants

The average age of the respondents was 33.7 years, with an SD of 12.9 (**Table 1**). The highest proportion of respondents was from the age group of 18 to 24 years (28.9%). About 47% of the respondents were women, while most of the respondents (86.9%) were Muslims. Nearly two-thirds of the respondents (61.6%) were married, while only 20.6% of the respondents had less than a secondary education level. About two-thirds of the respondents (64.3%) were from rural areas, while 31.9% were from the Dhaka division. One-third of the respondents were students and unemployed. The mean household members were 5.0, while the mean household income was 37627 Taka. Table 1 also shows that sample characteristics were almost nationally representative about age, sex, religion, marital status, place of residence, and the mean number of household members (Column 3, Table 1).

**Table 1:** Background characteristics of the study population.

| Variables | Study sample, n (%) | National population (%) |
| --- | --- | --- |
| **Age (in years)** |  |  |
| 18-24 | 432 (28.9) | 20.1 |
| 25-30 | 362 (24.2) | 19.7 |
| 31-39 | 254 (17.0) | 22.7 |
| 40-49 | 236 (15.8) | 18.5 |
| 50+ | 213 (14.2) | 19.1 |
| Mean (SD) | 33.7 (12.9) |  |
| **Sex** |  |  |
| Women | 692 (46.2) | 50.1 |
| Men | 805 (53.8) | 49.9 |
| **Religion** |  |  |
| Others | 196 (13.1) | 9.3 |
| Muslim | 1301 (86.9) | 90.7 |
| **Marital status** |  |  |
| Unmarried | 575 (38.4) | 34.8 |
| Married | 922 (61.6) | 65.2 |
| **Education** |  |  |
| No education | 129 (8.6) | 28.9 |
| Primary | 179 (12.0) | 27.5 |
| Secondary and higher secondary | 448 (29.9) | 43.6 |
| Graduate | 400 (26.7) |  |
| Masters and MPhil/PhD | 341 (22.8) |  |
| **Place of residence** |  |  |
| Rural area | 963 (64.3) | 65.0 |
| Urban area (other than city corporation) | 179 (12.0) | 35.0 |
| City Corporation | 355 (23.7) |  |
| **Administrative division of Bangladesh** |  |  |
| Barishal | 114 (7.6) | 5.7 |
| Chattogram | 253 (16.9) | 20.1 |
| Dhaka | 478 (31.9) | 25.1 |
| Khulna | 137 (9.2) | 10.8 |
| Mymensingh | 108 (7.2) | 7.8 |
| Rajshahi | 180 (12.0) | 12.7 |
| Rangpur | 114 (7.6) | 10.9 |
| Sylhet | 113 (7.5) | 6.8 |
| **Occupation** |  |  |
| Government, private, & NGO sector job | 202 (13.5) |  |
| Professional | 277 (18.5) |  |
| Homemakers | 348 (23.2) |  |
| Students and unemployed | 473 (31.6) |  |
| Agriculture and Day Laborer | 102 (6.8) |  |
| Others | 95 (6.3) |  |
| **Household members, Mean (SD)** | 5.0 (2.0) | 4.6 |
| **Household income, Mean (SD)** | 37627.2 (81295.9) |  |
| **Total** | 1497 (100.0) |  |

### 3.2. Prevalence of the COVID-19 Vaccine Hesitancy

About 43% of the respondents reported that they would surely receive the COVID-19 vaccine if available for free, while 17.7 percent would probably receive the vaccine. Besides, 22.8% of respondents mentioned that they would strongly encourage their family members to receive the vaccine, while 37.5% would encourage their family members to take the vaccine (**Fig. 1**). On the other hand, 12.3% of the respondents stated that they would delay receiving the vaccine, followed by 13.2% were unsure about what they would do, 7% would probably not receive it, and 6.9% would surely not receive the vaccine. Similarly, if the family or friends were thinking of receiving the COVID-19 vaccine, 16.8% of the respondents supported the statement that they would ask their family members or friends to delay in receiving the vaccine, while 16.9% would not say anything about it, 2.9% would discourage their family members and friends from receiving the vaccine, and 3% would forbid their family members and friends to receive the vaccine. Overall, the mean score of the hesitancy scale was 4.93 with an SD of 2.68. The accuracy of the hesitancy scale was 41.1% (4.93/12*100), showing that 41.1% of the respondents had hesitancy to receive the COVID-19 vaccine.

**Fig. 1.**
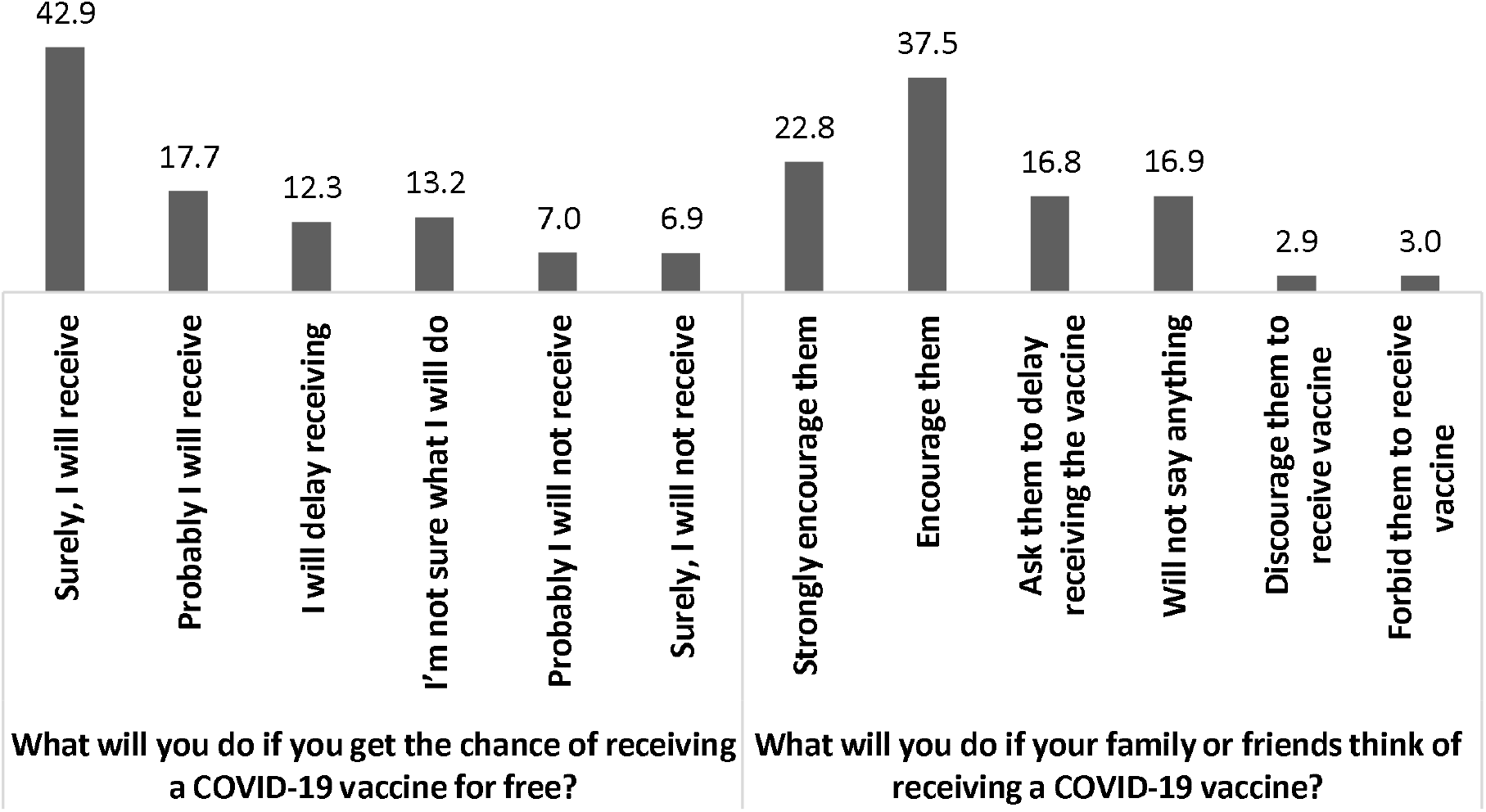
Prevalence (%) of COVID-19 vaccine hesitancy among the study population.

### 3.3. Vaccine Hesitancy by Respondents Background Characteristics

The mean level of hesitancy was statistically significantly (p < 0.05) varied by respondents’ sex, religion, place of residence, the administrative division of Bangladesh (Table 2). The mean hesitancy was higher among women, Muslims, respondents from city corporation areas and Khulna division.

**Table 2:**
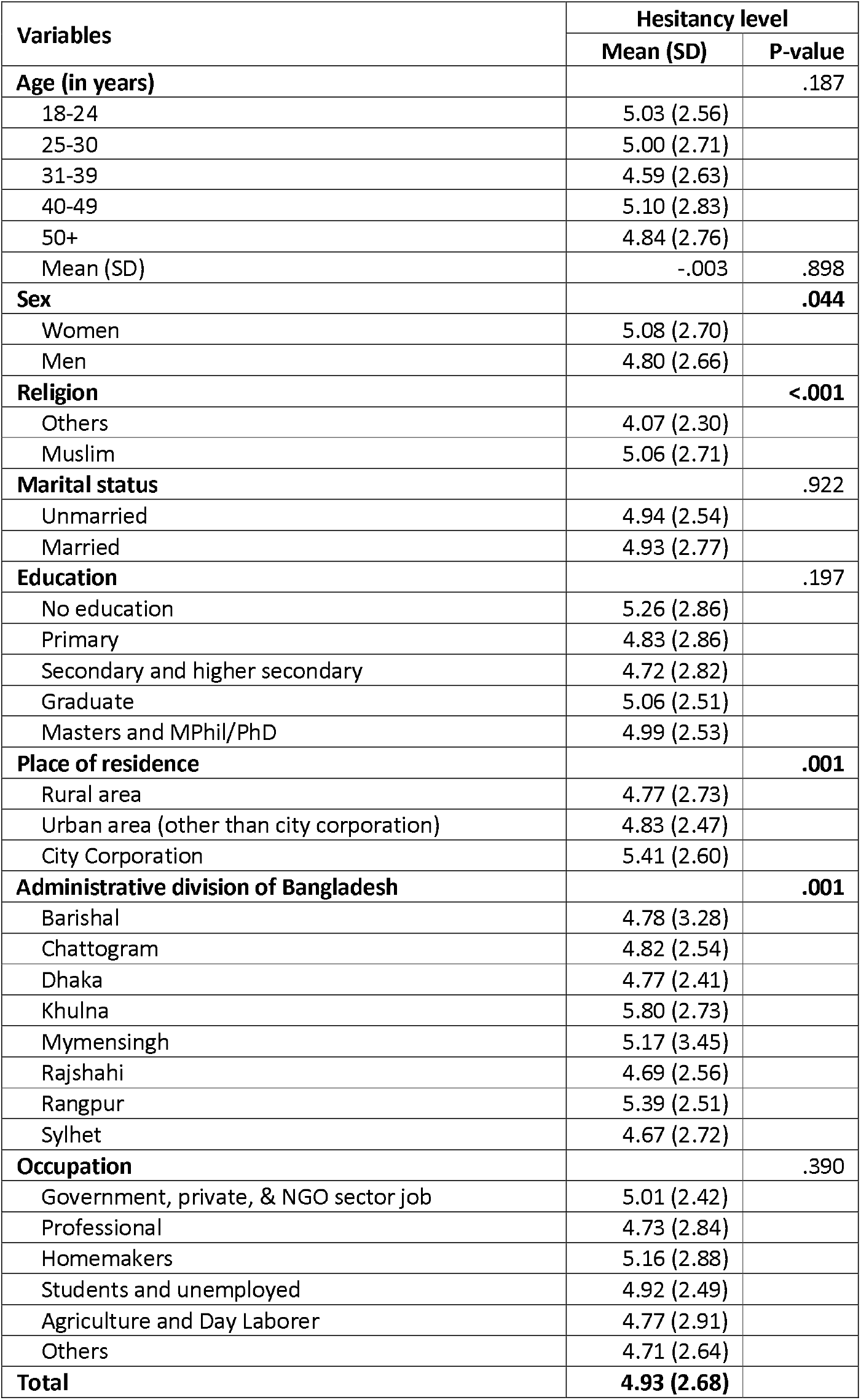
Mean level of COVID-19 vaccine hesitancy by the respondent’s characteristics.

| Variables | Hesitancy level |  |
| --- | --- | --- |
|  | Mean (SD) | P-value |
| **Age (in years)** |  | .187 |
| 18-24 | 5.03 (2.56) |  |
| 25-30 | 5.00 (2.71) |  |
| 31-39 | 4.59 (2.63) |  |
| 40-49 | 5.10 (2.83) |  |
| 50+ | 4.84 (2.76) |  |
| Mean (SD) | -.003 | .898 |
| **Sex** |  | **.044** |
| Women | 5.08 (2.70) |  |
| Men | 4.80 (2.66) |  |
| **Religion** |  | **<.001** |
| Others | 4.07 (2.30) |  |
| Muslim | 5.06 (2.71) |  |
| **Marital status** |  | .922 |
| Unmarried | 4.94 (2.54) |  |
| Married | 4.93 (2.77) |  |
| **Education** |  | .197 |
| No education | 5.26 (2.86) |  |
| Primary | 4.83 (2.86) |  |
| Secondary and higher secondary | 4.72 (2.82) |  |
| Graduate | 5.06 (2.51) |  |
| Masters and MPhil/PhD | 4.99 (2.53) |  |
| **Place of residence** |  | **.001** |
| Rural area | 4.77 (2.73) |  |
| Urban area (other than city corporation) | 4.83 (2.47) |  |
| City Corporation | 5.41 (2.60) |  |
| **Administrative division of Bangladesh** |  | **.001** |
| Barishal | 4.78 (3.28) |  |
| Chattogram | 4.82 (2.54) |  |
| Dhaka | 4.77 (2.41) |  |
| Khulna | 5.80 (2.73) |  |
| Mymensingh | 5.17 (3.45) |  |
| Rajshahi | 4.69 (2.56) |  |
| Rangpur | 5.39 (2.51) |  |
| Sylhet | 4.67 (2.72) |  |
| **Occupation** |  | .390 |
| Government, private, & NGO sector job | 5.01 (2.42) |  |
| Professional | 4.73 (2.84) |  |
| Homemakers | 5.16 (2.88) |  |
| Students and unemployed | 4.92 (2.49) |  |
| Agriculture and Day Laborer | 4.77 (2.91) |  |
| Others | 4.71 (2.64) |  |
| **Total** | **4.93 (2.68)** |  |

### 3.4. Vaccine Hesitancy by Behavioral Practices to Prevent COVID-19

The mean level of vaccine hesitancy significantly varied by the participants’ behavioral practices to prevent COVID-19 (Fig.2). The respondents who never wore the mask were more hesitant than others. Similarly, those who never avoided crowds were also more hesitant.

**Fig. 2.**
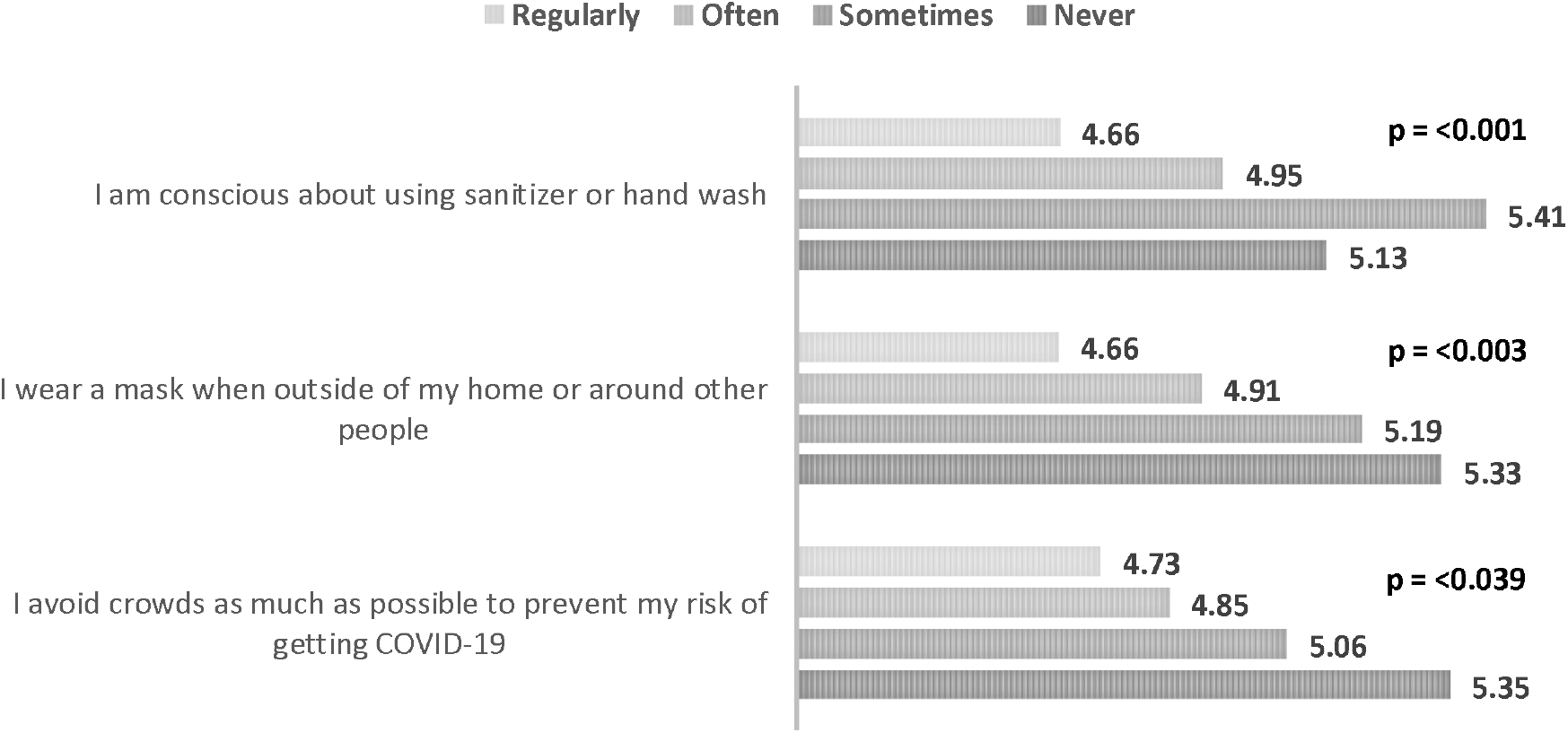
Mean Level of Vaccine Hesitancy* by Behavioral Practices to Prevent COVID-19. *A score above 4 shows vaccine hesitancy.

### 3.5. Vaccine Hesitancy by Knowledge about the COVID-19 Vaccine and Vaccination Process

**Fig.3** illustrates the level of vaccine hesitancy by knowledge about the COVID-19 vaccine. The respondents who strongly disagreed with the statement that the COVID-19 vaccine has very mild side effects were more hesitant to receive the vaccine. The participants who had incorrect knowledge about the vaccination process were more hesitant than those who had correct knowledge (**Fig. 4**).

**Fig. 3.**
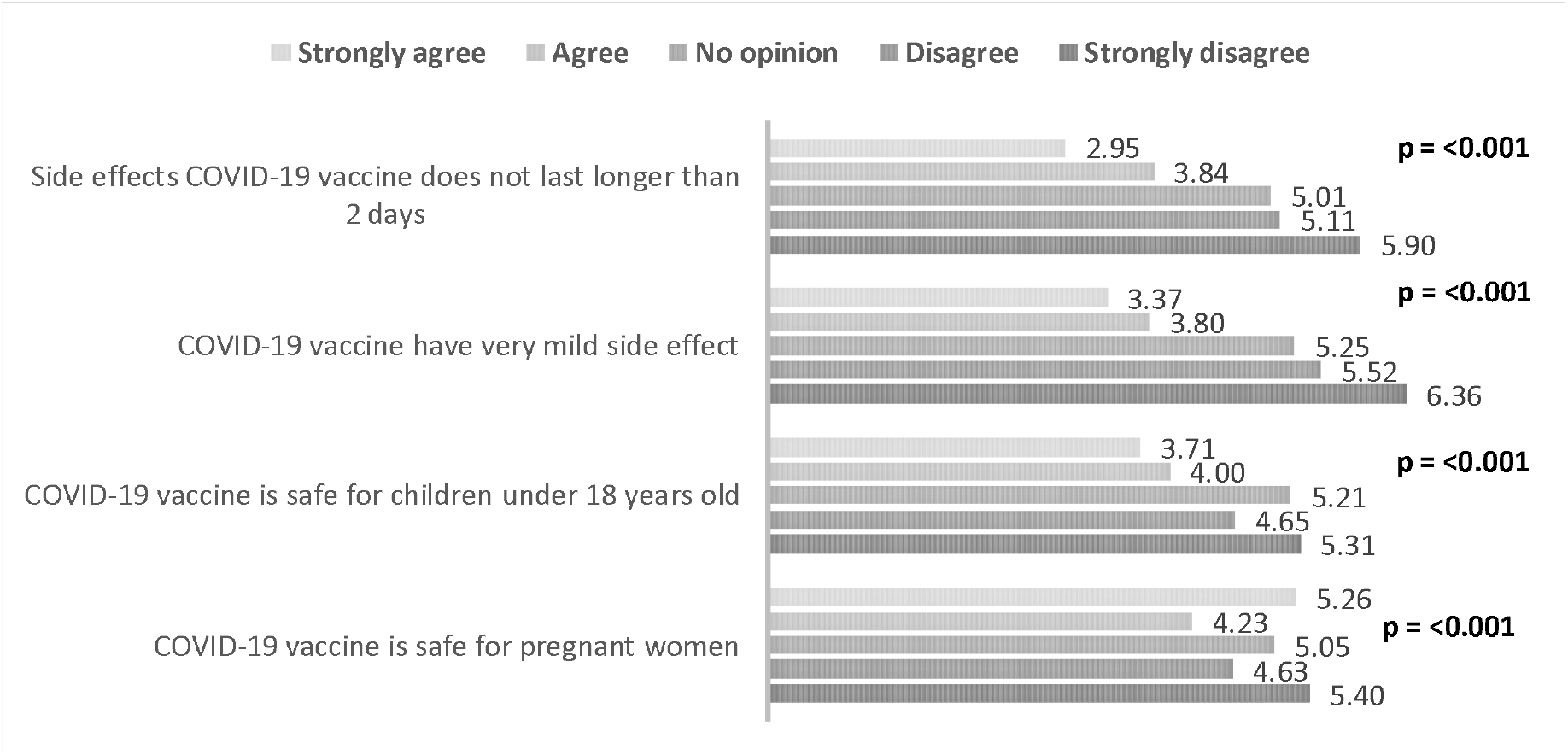
Mean Level of Vaccine Hesitancy* by Knowledge about the COVID-19 Vaccine. *A score above 4 shows vaccine hesitancy.

**Fig. 4.**
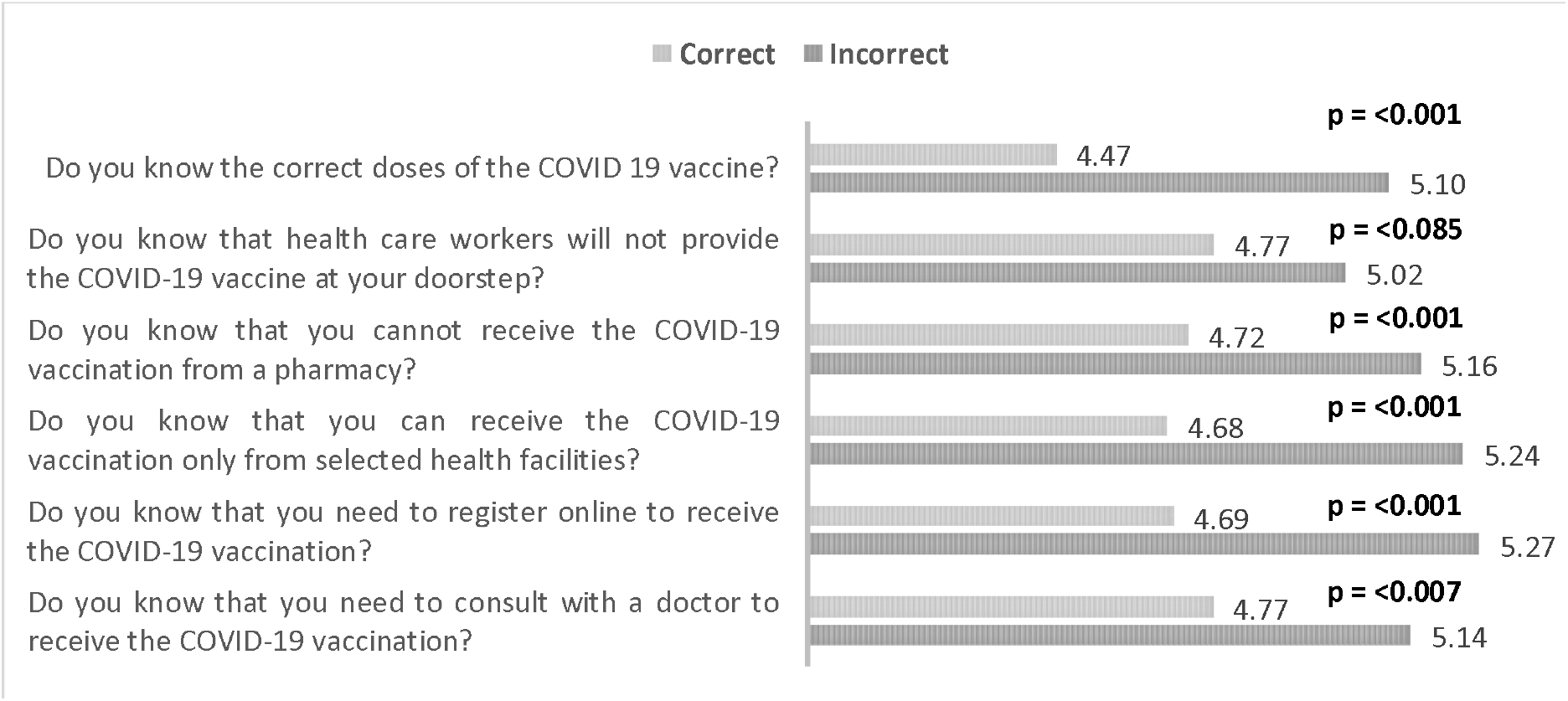
Mean Level of Vaccine Hesitancy* by Knowledge about the COVID-19 Vaccination Process. *A score above 4 shows vaccine hesitancy.

### 3.6. Vaccine Hesitancy by Attitude towards COVID-19 Vaccine and Vaccine Conspiracy

Table 3 depicts the level of hesitancy by attitudes towards the COVID-19 vaccine and conspiracy beliefs associated with it. The respondents who had more negative attitudes and conspiracy beliefs had more hesitancy to accept the COVID-19 vaccine. The respondents who did not trust the COVID-19 vaccine and believed that the vaccine would probably not work had more hesitancy. On the other hand, the respondents who strongly agreed to the notion that the Coronavirus is a myth to force vaccinations on people had more hesitancy. In contrast, the respondents who strongly disagreed with the statement that the COVID-19 vaccines made in America and Europe are safer than those made in other countries, and the COVID-19 vaccines made in India are safer than those made in other countries had more hesitancy.

**Table 3:** Mean Level of Vaccine Hesitancy by Attitude and Conspiracy towards COVID-19 Vaccine.

| Variables and Items | Mean level of hesitancy* |  |  |  |  | p-value |
| --- | --- | --- | --- | --- | --- | --- |
|  | Strongly disagree | Disagree | No opinion | Agree | Strongly agree |  |
| **Attitude towards COVID-19 Vaccine** |  |  |  |  |  |  |
| I think the COVID-19 vaccine probably will not work | 3.49 | 3.91 | 5.28 | 5.72 | 6.61 | <.001 |
| I do not trust the COVID-19 vaccine | 3.71 | 4.36 | 5.88 | 6.37 | 6.54 | <.001 |
| I think the COVID-19 vaccine is unnecessary | 3.86 | 4.71 | 6.10 | 6.80 | 6.70 | <.001 |
| I think it is not important to get a vaccine to protect people from the COVID-19** | 5.47 | 6.67 | 5.92 | 4.73 | 3.60 | <.001 |
| I do not need a COVID-19 vaccine because I am healthy and at low risk for infection | 3.54 | 4.04 | 5.65 | 5.93 | 6.52 | <.001 |
| I do not need a COVID-19 vaccine because even if I get infected, I will not become seriously ill | 3.71 | 4.12 | 5.56 | 6.00 | 6.59 | <.001 |
| **Covid-19 Vaccine Conspiracy** |  |  |  |  |  |  |
| Pharmaceutical companies are encouraging the spread of Coronavirus to make a profit through selling vaccine | 3.83 | 4.77 | 5.42 | 5.35 | 6.40 | <.001 |
| The Coronavirus is a myth to force vaccinations on people | 4.03 | 4.99 | 5.65 | 5.52 | 7.15 | <.001 |
| Drug companies cover up the side effects of vaccines | 3.32 | 4.29 | 5.23 | 5.81 | 6.68 | <.001 |
| People are deceived about the effectiveness of vaccines | 3.27 | 4.34 | 5.33 | 6.00 | 6.41 | <.001 |
| COVID-19 vaccine can result into autism | 3.85 | 4.29 | 5.33 | 5.48 | 5.25 | <.001 |
| A coronavirus vaccination could give one coronavirus | 3.64 | 4.44 | 5.44 | 5.65 | 5.88 | <.001 |
| COVID-19 vaccines made in America and Europe are safer than those made in other countries** | 5.86 | 5.12 | 5.20 | 4.65 | 3.86 | <.001 |
| COVID-19 vaccines made in China and Russia are safer than those made in other countries** | 5.02 | 4.78 | 5.13 | 4.46 | 4.09 | 0.001 |
| COVID-19 vaccines made in India are safer than those made in other countries** | 6.15 | 4.89 | 4.95 | 3.75 | 3.27 | <.001 |

\*A score above 4 shows vaccine hesitancy; \*\* Items were reverse coded.

### 3.7. Vaccine Hesitancy by the Constructs of Health Belief Model

The mean level of vaccine hesitancy by the health belief model components is presented in Table 4. The respondents who strongly disagreed with the statements related to perceived susceptibility, severity, and benefits were more hesitant to receive a vaccine. For example, the respondents who strongly disagreed with the statement that I think the complications of the COVID-19 will decrease if I get vaccinated and then get infected with the Coronavirus had more hesitancy. On the other hand, the respondents who strongly agreed with the statements related to perceived barriers were also more hesitant to receive the COVID-19 vaccine. For instance, the respondents who strongly agreed that registering for COVID-19 vaccination is difficult for them had more hesitancy.

**Table 4:** Mean Level of Vaccine Hesitancy by Health Belief Model related to COVID-19 vaccine.

| Health Belief Model | Mean level of hesitancy* |  |  |  |  | p-value |
| --- | --- | --- | --- | --- | --- | --- |
|  | Strongly disagree | Disagree | No opinion | Agree | Strongly agree |  |
| **Perceived Susceptibility** |  |  |  |  |  |  |
| I am worried about the likelihood of getting infected by COVID-19 | 6.24 | 5.09 | 5.53 | 4.42 | 4.21 | <.001 |
| I am at high risk of COVID-19 because of my health conditions | 5.57 | 4.83 | 5.06 | 4.43 | 4.46 | <.001 |
| **Perceived Severity** |  |  |  |  |  |  |
| I will be very sick if I get infected by COVID-19 | 6.33 | 5.04 | 5.27 | 4.30 | 3.48 | <.001 |
| I am very concerned that I could die from COVID-19 | 5.99 | 4.85 | 5.05 | 4.52 | 3.71 | <.001 |
| **Perceived Benefits** |  |  |  |  |  |  |
| I think vaccination is good because it will make me less worried about COVID-19 | 6.69 | 6.25 | 5.95 | 4.44 | 3.30 | <.001 |
| I believe vaccination will decrease my risk of getting infected by COVID-19 | 6.97 | 6.42 | 5.78 | 4.38 | 3.28 | <.001 |
| I think the complications of COVID-19 will decrease if I get vaccinated and then get infected with the Coronavirus. | 7.37 | 6.23 | 5.50 | 4.15 | 3.37 | <.001 |
| **Perceived Barriers** |  |  |  |  |  |  |
| I am worried that the possible side effects of the COVID-19 vaccination would interfere with my usual activities | 3.74 | 3.92 | 5.47 | 4.76 | 5.78 | <.001 |
| I am concerned about the efficacy of the COVID-19 vaccine | 4.16 | 4.16 | 5.15 | 4.99 | 5.78 | <.001 |
| I have a concern that I may receive faulty/fake COVID-19 vaccine | 3.12 | 4.06 | 4.98 | 5.04 | 5.93 | <.001 |
| It concerns me that the development of a COVID-19 vaccine is too rushed to test its safety properly | 2.97 | 3.90 | 5.22 | 5.53 | 6.48 | <.001 |
| I am concerned about the long-term side effects of the COVID-19 vaccination | 3.25 | 3.91 | 5.17 | 4.98 | 6.04 | <.001 |
| Registering for COVID-19 vaccination is difficult for me | 3.77 | 4.59 | 5.49 | 5.16 | 5.92 | <.001 |

\*A score above 4 shows vaccine hesitancy.

### 3.8. Predictors of COVID-19 Vaccine Hesitancy

The independent variables significant at the bivariate level were then entered into the multiple linear regression model after checking the assumptions and multicollinearity (**Table 5**). We produced three models. The first model included the socio-economic and demographic characteristics of the study population, while the second model included all the variables of model 1 plus knowledge, attitudes, conspiracy, and behavioral practices related to the COVID-19 vaccine. The third model included all the variables of model 2 plus all the components of HBM. All the regression models were highly significant. The adjusted R^2^ of the final regression model (model 3) was 0.379. Moreover, compared to model 1, successive models had higher R^2^, and lower AIC (Akaike Information Criterion) and BIC (Bayesian Information Criterion) showed better model fitting.

**Table 5:** Factors affecting level COVID-19 vaccine of hesitancy among adult population in Bangladesh.

| Independent variables | Model 1 |  | Model 2 |  | Model 3 |  |
| --- | --- | --- | --- | --- | --- | --- |
|  | Beta (SE) | p | Beta (SE) | p | Beta (SE) | p |
| **Sex (Women as RC)** |  |  |  |  |  |  |
| Men | -.055 (.136) | .031 | -.052 (.117) | .016 | -.046 (.113) | .030 |
| **Religion (Others as RC)** |  |  |  |  |  |  |
| Muslim | .129 (.208) | <.001 | .081 (.176) | <.001 | .057 (.172) | .009 |
| **Place of residence (Rural as RC)** |  |  |  |  |  |  |
| Urban area (other than city corporation) | -.013 (.215) | .613 | .045 (.190) | .049 | .042 (.184) | .063 |
| City Corporation | .138 (.173) | <.001 | .136 (.154) | <.001 | .132 (.152) | <.001 |
| **Administrative division of Bangladesh (Sylhet as RC)** |  |  |  |  |  |  |
| Barishal | .001 (.350) | .988 | .055 (.294) | .058 | .060 (.286) | .035 |
| Chattogram | .037 (.299) | .375 | .046 (.250) | .189 | .061 (.242) | .069 |
| Dhaka | -.016 (.279) | .749 | .031 (.234) | .448 | .020 (.227) | .604 |
| Khulna | .126 (.334) | <.001 | .086 (.280) | .004 | .093 (.271) | .001 |
| Mymensingh | .055 (.353) | .104 | .076 (.297) | .008 | .073 (.288) | .008 |
| Rajshahi | .014 (.315) | .721 | .002 (.264) | .942 | .015 (.256) | .629 |
| Rangpur | .113 (.355) | .001 | .070 (.298) | .018 | .068 (.288) | .017 |
| **Behavioral practice to prevent COVID-19** |  |  | .010 (.023) | .658 | .020 (.023) | .389 |
| **Knowledge about the COVID-19 vaccine** |  |  | -.103 (.027) | <.001 | -.072 (.026) | .001 |
| **Knowledge about the vaccination process** |  |  | -.058 (.034) | .020 | -.058 (.033) | .018 |
| **Covid-19 vaccine conspiracy** |  |  | .184 (.014) | <.001 | .105 (.015) | <.001 |
| **Attitude towards COVID-19 vaccine** |  |  | .398 (.017) | <.001 | .291 (.018) | <.001 |
| **Health Belief Model** |  |  |  |  |  |  |
| Perceived susceptibility |  |  |  |  | -.020 (.037) | .418 |
| Perceived severity |  |  |  |  | -.079 (.039) | .002 |
| Perceived benefits |  |  |  |  | -.180 (.039) | <.001 |
| Perceived barriers |  |  |  |  | .180 (.019) | <.001 |
| **Model Summary** |  |  |  |  |  |  |
| N | 1497 |  | 1497 |  | 1497 |  |
| R Square | .052 |  | .341 |  | .387 |  |
| Adjusted R Square | .045 |  | .334 |  | .379 |  |
| R Square Change | - |  | .289 |  | .046 |  |
| AIC | 2898.62 |  | 2364.59 |  | 2263.57 |  |
| BIC | 2962.35 |  | 2454.88 |  | 2375.11 |  |

Beta: Standardized coefficient, SE: Standard error, P= Probability value, RC = Reference category.

According to model 3, men had less hesitancy (β = -0.046, p = 0.030) than women. The Muslims (β = 0.057, p = 0.009) and the respondents of city corporation areas (β = 0.132, p <0.001) had more hesitancy than that of others. Compared to the Sylhet division, the participants from Barishal (β = 0.060, p =0.035), Khulna (β = 0.093, p =0.001), Mymensingh (β = 0.073, p =0.008), and Rangpur (β = 0.068, p =0.017) had higher hesitancy. With increasing the knowledge about vaccine (β = -0.072, p=0.001) and knowledge about vaccination process (β= -0.058, p = 0.018), hesitancy tended to decrease. On the other hand, with increasing negative attitudes towards vaccine (β = 0.291, p <0.001) and conspiracy beliefs towards vaccine (β = 0.105, p=0.004), the hesitancy increased. The perceived severity (β = -0.079, p=0.002) and benefits of COVID-19 (β = -0.180, p <0.001) reduced the hesitancy, while perceived barriers of COVID-19 vaccination (β = 0.180, p <0.001) increased the hesitancy.

## 4. Discussions

The study found that about 14% of the respondents have asserted their intention towards not receiving the vaccines, while 16.8% of the respondents have reported that they will suggest their friends and families delaying receiving COVID-19 vaccines. The study also found that 2.9% of the respondents will discourage their family members from receiving the vaccination, and 3% of the respondents will forbid family members to do so. Overall, this study found a 41.1% hesitancy to receive the COVID-19 vaccine, which is higher than the only available earlier study (32.5%) conducted in Bangladesh [14]. Perhaps, the difference can be explained by the methodological differences as the earlier study collected data online while our study collected data using both online and face-face-to-face interviews.

This study found that women were more hesitant than men regarding receiving the COVID-19 vaccination, which is supported by the findings of other studies [23,24]. The low economic ability of women [25], women’s concerns regarding the potentially harmful effect of the COVID-19 vaccines towards developing baby in the womb, and young children [26], conspiracy beliefs among women regarding COVID-19 vaccine imposed subfertility, infertility, and miscarriages [27], and less perceived susceptibility regarding contracting Coronavirus among women [28] are playing a role towards constructing increased vaccine hesitancy among the women.

Our study shows that religion was significantly associated with vaccine hesitancy, which is in line with different other studies from both the COVID-19 and non-COVID-19 contexts [29,30]. The Muslims had more hesitancy about the receipt of coronavirus vaccination in the current study. The notion of considering vaccines as ‘medical assault’, doubts regarding the ingredients of the vaccines (doubts over the inclusion of ingredients like pork gelatin) may play a role behind the increased hesitancy of Muslim people regarding COVID-19 vaccines [31]. Considering the COVID-19 vaccines as a ‘western plot’ to sterilize Muslim women is another way of thinking explored within the Muslim communities for which this vaccine is largely being discouraged by the community [32]. In different earlier non-COVID-19 context too, such as in the cases of measles, mumps, and rubella (MMR), religious ruling against vaccines considering them as ‘*haram*’ (forbidden) due to the suspected presence of ingredients derived from pigs, receiving vaccines were discouraged mainly [33]. Religious fatalism among the Muslims, including the beliefs that ‘everything is in the hands of Allah,’ perception of the powerlessness concerning health condition, and sense of inability of avoiding death when it is the will of Allah, influences the perception of health among Muslims [34] and such perspectives on health, in this case, is possibly growing vaccine hesitancy among the Muslims [35].

The findings of this study show that respondents from the city corporation areas are more hesitant about the uptake of the COVID-19 vaccines. Due to having more exposure to the different online and offline sources of information, the residents of the city corporation have more possibility of producing fear-driven stigma and conspiracy beliefs regarding COVID-19 vaccines, which may explain their higher level of vaccine hesitancy. In a non-COVID-19 context (dengue vaccine), the broader access towards negative media information in urban areas regarding vaccines has been found responsible for a high level of vaccine hesitancy [36]. However, studies on the COVID-19 context have also found that rural inhabitants are more likely to experience vaccine-related hesitancy [24] than the urban counterparts.

Our study shows that with the increased level of knowledge about the COVID-19 vaccine and its associated processes, the hesitancy decreased. The vaccine-related knowledge, which creates awareness regarding vaccine’s role in decreasing the risks of the diseases among individuals, plays a role in lessening vaccine hesitancy [37]. Being knowledgeable about the vaccination process has been found in other studies too as a significant predictor of vaccine hesitancy [37,38], leaving ample scopes for vaccine promotion strategies targeting the information gap.

Attitudes toward the COVID-19 vaccine have been appeared to be one of the strongest predictors of vaccine hesitancy. The negative attitudes towards the COVID-19 vaccine, including perceiving less importance of vaccines, mistrust about effectiveness, were associated with increasing vaccine hesitancy among the respondents of this study. A negative or anti-COVID-19 vaccination attitude is formed because of the low confidence in vaccine safety [39] and vaccine benefits [38], concerns regarding potential side effects [20], and also the newness of the vaccine [12]. The finding of this current study is in line with other studies, where it was shown that people having a more negative attitude towards the COVID-19 vaccines are less willing to receive the vaccine [38]

The conspiracy about the COVID-19 vaccines, vaccination process, pharmaceutical companies’ roles, vaccine manufacturers, and consequences have been responsible for increasing the vaccine hesitancy in our study. In various other studies, conspiracy narratives have been regarded as the seed bearer of vaccine hesitancy and considered responsible for resistance against the COVID-19 vaccination programs [15]. The hesitancy towards receiving the COVID-19 vaccines has been seen to be significantly influenced by the presence of different conspiracy beliefs [17]. Various conspiracies, including misinformation regarding the origin of the COVID-19, COVID-19 vaccines trials [40], suspicions around vaccine manufacturers (pharmaceuticals companies and country of origin) [17,41] regarding vaccine efficacy and safety have been considered as responsible in other studies for fueling pre-existing fears, fostering mistrust, doubts, and cynicism over new vaccines, and lowering the COVID-19 vaccine uptake intention of mass people [42].

The study used the constructs of HBM as independent variables to predict vaccine hesitancy, and it was found that all the components of HBM except perceived susceptibility were strongly predicting the level of vaccine hesitancy among individuals. Our study found a strong negative association between perceived severity and the COVID-19 vaccine hesitancy. Evidence shows that perceiving less severity of disease lowers the sense of threat among individuals, which ultimately grows hesitancy regarding the necessity of receiving vaccines as a preventive behavior [43]. A similar association has been observed in other studies, too [9,44]. Our study also found that perceived benefits are negatively associated with vaccine hesitancy. Considering a particular action (in this case, receiving vaccination) as effective in preventing a disease, which is perceived benefits according to HBM constructs, motivates individuals in adopting the behaviors [45]. Perceived barriers were positively associated with vaccine hesitancy in our study. Different perceived structural and attitudinal barriers have been found in other studies [46] as responsible for the vaccine hesitancy, such as lack of information about the vaccination and its adverse effects [47], not getting access to the vaccination coverage [48], affordability issues [49], individual’s negative concerns regarding side effects and efficacy of the vaccine [9].

Though this study explored the extent of hesitancy and associated factors about the COVID-19 vaccine in Bangladesh, which will help policymakers develop messages to counter the anti-vaccine campaign, some limitations should be considered in interpreting the results. This study used a non-probability sampling; therefore, we should be careful about the generalization of the findings. Though this study tried to represent the national population in terms of age, sex, residence, region, marital status, and religion, the distribution of education among the respondents is to some extent not comparable to national data. This study collected self-reported data that may suffer from reporting bias to some extent. Finally, this research used a cross-sectional study design which limits the causality.

## 5. Conclusions

The findings of this study suggest that a vigorous campaign should be developed to reduce the negative attitude towards the COVID-19 vaccine and conspiracy against the COVID-19 vaccine and expand population-wide faith concerning vaccination. The authorities concerned with importing the COVID-19 vaccines in Bangladesh should intervene to contract and engage more than one vaccine manufacturing organization to avoid skepticism toward any single country of origin or pharmaceutical company. The policymakers should also think about revisiting the policy of the online registration process to receive the COVID-19 vaccine as many people cannot register themselves for receiving vaccination due to the digital divide in the country. The digital divide prevents the population from the lower socio-economic strata and women from receiving the COVID-19 vaccine. Thus, initiatives like text message service using the mobile phone operator can be introduced.

## Data Availability

Data are available from the Corresponding Author.

## Acknowledgments

We would like to thank the participants of this study. We would also like to thank the data collectors for their contribution amidst this challenging time of the COVID-19 pandemic.

## References

[1] Larson HJ, Jarrett C, Eckersberger E, Smith DMD, Paterson P. Understanding vaccine hesitancy around vaccines and vaccination from a global perspective: A systematic review of published literature, 2007-2012. Vaccine 2014;32:2150–9. https://doi.org/10.1016/j.vaccine.2014.01.081.

[2] Wiysonge CS, Ndwandwe D, Ryan J, Jaca A, Batouré O, Anya BPM, et al. Vaccine hesitancy in the era of COVID-19: Could lessons from the past help in divining the future? Hum Vaccines Immunother 2021. https://doi.org/10.1080/21645515.2021.1893062.

[3] MacDonald NE, Eskola J, Liang X, Chaudhuri M, Dube E, Gellin B, et al. Vaccine hesitancy: Definition, scope and determinants. Vaccine 2015;33:4161–4. https://doi.org/10.1016/j.vaccine.2015.04.036.

[4] Robinson E, Jones A, Lesser I, Daly M. International estimates of intended uptake and refusal of COVID-19 vaccines: A rapid systematic review and meta-analysis of large nationally representative samples. Vaccine 2021;39:2024–34. https://doi.org/10.1016/j.vaccine.2021.02.005.

[5] Saad-Roy CM, Wagner CE, Baker RE, Morris SE, Farrar J, Graham AL, et al. Immune life history, vaccination, and the dynamics of SARS-CoV-2 over the next 5 years. Science (80-) 2020;370:811–8https://doi.org/10.1126/science.abd7343.

[6] United Nations Bangladesh. COVID-19 quarterly report: Supporting the government response to the pandemic. 2020.

[7] United Nations, Department of Economic and Social Affairs PD. World Population Prospects 2019, custom data acquired via website 2020. https://population.un.org/wpp/DataQuery/ (accessed September 14, 2020).

[8] Directorate General of Health Services (DGHS). COVID-19 vaccination dashboard, 2021. 2021. http://103.247.238.92/webportal/pages/covid19-vaccination.php (accessed April 19, 2021).

[9] Lin Y, Hu Z, Zhao Q, Alias H, Danaee M, Wong LP. Understanding COVID-19 vaccine demand and hesitancy: A nationwide online survey in China. PLoS Negl Trop Dis 2020;14:e0008961. https://doi.org/10.1371/journal.pntd.0008961.

[10] Mercadante AR, Law A V. Will they, or Won’t they? Examining patients’ vaccine intention for flu and COVID-19 using the Health Belief Model. Res Soc Adm Pharm 2021. https://doi.org/10.1016/j.sapharm.2020.12.012.

[11] Ruiz JB, Bell RA. Predictors of intention to vaccinate against COVID-19: Results of a nationwide survey. Vaccine 2021;39:1080–6. https://doi.org/10.1016/j.vaccine.2021.01.010.

[12] Paul E, Steptoe A, Fancourt D. Anti-vaccine attitudes and risk factors for not agreeing to vaccination against COVID-19 amongst 32,361 UK adults: Implications for public health communications. MedRxiv 2020. https://doi.org/10.1101/2020.10.21.20216218.

[13] Soares P, Rocha JV, Moniz M, Gama A, Laires PA, Pedro AR, et al. Factors associated with COVID-19 vaccine hesitancy. Vaccines 2021;9. https://doi.org/10.3390/vaccines9030300.

[14] Ali M, Hossain A. What is the extent of COVID-19 vaccine hesitancy in Bangladesh?1: A cross-sectional rapid national survey. MedRxiv 2021. https://doi.org/10.1101/2021.02.17.21251917.

[15] Khan YH, Mallhi TH, Alotaibi NH, Alzarea AI, Alanazi AS, Tanveer N, et al. Threat of COVID-19 vaccine hesitancy in Pakistan: The need for measures to neutralize misleading narratives. Am J Trop Med Hyg 2020;103:603–4. https://doi.org/10.4269/ajtmh.20-0654.

[16] Dubé E, Vivion M, MacDonald NE. Vaccine hesitancy, vaccine refusal and the anti-vaccine movement: Influence, impact and implications. Expert Rev Vaccines 2014;14:99–117. https://doi.org/10.1586/14760584.2015.964212.

[17] Sallam M, Dababseh D, Eid H, Al-Mahzoum K, Al-Haidar A, Taim D, et al. High rates of covid-19 vaccine hesitancy and its association with conspiracy beliefs: A study in jordan and kuwait among other arab countries. Vaccines 2021;9:1–16. https://doi.org/10.3390/vaccines9010042.

[18] Latkin CA, Dayton L, Yi G, Colon B, Kong X. Mask usage, social distancing, racial, and gender correlates of COVID-19 vaccine intentions among adults in the US. PLoS One 2021;16. https://doi.org/10.1371/journal.pone.0246970.

[19] Momplaisir F, Haynes N, Nkwihoreze H, Nelson M, Werner RM, Jemmott J. Understanding drivers of coronavirus disease 2019 vaccine hesitancy among Blacks. Clin Infect Dis 2021. https://doi.org/10.1093/cid/ciab102.

[20] Neumann-Böhme S, Varghese NE, Sabat I, Barros PP, Brouwer W, van Exel J, et al. Once we have it, will we use it? A European survey on willingness to be vaccinated against COVID-19. Eur J Heal Econ 2020;21:977–82. https://doi.org/10.1007/s10198-020-01208-6.

[21] Rhodes A, Hoq M, Measey MA, Danchin M. Intention to vaccinate against COVID-19 in Australia. Lancet Infect Dis 2020. https://doi.org/10.1016/S1473-3099(20)30724-6.

[22] Hossain MB, Alam MZ, Islam MS, Sultan S, Faysal MM, Rima S, et al. Data on COVID-19 vaccine hesitancy among the adult population in Bangladesh. Mendeley Data 2021;V1. https://doi.org/10.17632/prgh4bb3yf.1.

[23] Edwards B, Biddle N, Gray M, Sollis K. COVID-19 vaccine hesitancy and resistance: Correlates in a nationally representative longitudinal survey of the Australian population. PLoS One 2021. https://doi.org/10.1371/journal.pone.0248892.

[24] Khubchandani J, Sharma S, Price JH, Wiblishauser MJ, Sharma M, Webb FJ. COVID-19 vaccination hesitancy in the United States: A rapid national assessment. J Community Health 2021;46:270–7. https://doi.org/10.1007/s10900-020-00958-x.

[25] Chang WH. A review of vaccine effects on women in light of the COVID-19 pandemic. Taiwan J Obstet Gynecol 2020;59:812–20. https://doi.org/10.1016/j.tjog.2020.09.006.

[26] Skjefte M, Ngirbabul M, Akeju O, Escudero D, Hernandez-Diaz S, Wyszynski DF, et al. COVID-19 vaccine acceptance among pregnant women and mothers of young children: results of a survey in 16 countries. Eur J Epidemiol 2021. https://doi.org/10.1007/s10654-021-00728-6.

[27] Murewanhema G. Vaccination hesitancy among women of reproductive age in resource-challenged settings: A cause for public health concern. Pan Afr Med J 2021;38. https://doi.org/10.11604/pamj.2021.38.336.28953.

[28] Jahangiry L, Bakhtari F, Sohrabi Z, Reihani P, Samei S, Ponnet K, et al. Risk perception related to COVID-19 among the Iranian general population: An application of the extended parallel process model. BMC Public Health 2020;20. https://doi.org/10.1186/s12889-020-09681-7.

[29] Walker TY, Elam-Evans LD, Singleton JA, Yankey D, Markowitz LE, Fredua B, et al. National, regional, state, and selected local area vaccination coverage among adolescents aged 13–17 years — United States, 2016. MMWR Morb Mortal Wkly Rep 2017;66:874–82. https://doi.org/10.15585/mmwr.mm6633a2.

[30] Olagoke AA, Olagoke OO, Hughes AM. Intention to vaccinate against the novel 2019 coronavirus disease: The role of health locus of control and religiosity. J Relig Health 2021;60:65–80. https://doi.org/10.1007/s10943-020-01090-9.

[31] Mahdi S, Ghannam O, Watson S, Padela AI. Predictors of physician recommendation for ethically controversial medical procedures: Findings from an exploratory national survey of American Muslim physicians. J Relig Health 2016;55:403–21. https://doi.org/10.1007/s10943-015-0154-y.

[32] Ali I. The COVID-19 pandemic: Making sense of rumor and fear. Med Anthropol 2020;39:376–9. https://doi.org/10.1080/01459740.2020.1745481.

[33] Loomba S, de Figueiredo A, Piatek SJ, de Graaf K, Larson HJ. Measuring the impact of exposure to COVID-19 vaccine misinformation on vaccine ntent in the UK and US. MedRxiv 2020. https://doi.org/10.1101/2020.10.22.20217513.

[34] Elbarazi I, Devlin NJ, Katsaiti MS, Papadimitropoulos EA, Shah KK, Blair I. The effect of religion on the perception of health states among adults in the United Arab Emirates: A qualitative study. BMJ Open 2017;7. https://doi.org/10.1136/bmjopen-2017-016969.

[35] Peretti-Watel P, Larson HJ, Ward JK, Schulz WS, Verger P. Vaccine hesitancy: Clarifying a theoretical framework for an ambiguous notion. PLoS Curr 2015;7. https://doi.org/10.1371/currents.outbreaks.6844c80ff9f5b273f34c91f71b7fc289.

[36] Migriño J, Gayados B, Birol KRJ, De Jesus L, Lopez CW, Mercado WC, et al. Factors affecting vaccine hesitancy among families with children 2 years old and younger in two urban communities in Manila, Philippines. West Pacific Surveill Response J WPSAR 2020;11:20–6. https://doi.org/10.5365/wpsar.2019.10.2.006.

[37] Eitze S, Heinemeier D, Schmid-Küpke NK, Betsch C. Decreasing vaccine hesitancy with extended health knowledge: Evidence from a longitudinal randomized controlled trial. Health Psychol 2021;40:77–88. https://doi.org/10.1037/hea0001045.

[38] Paul E, Steptoe A, Fancourt D. Attitudes towards vaccines and intention to vaccinate against COVID-19: Implications for public health communications. Lancet Reg Heal - Eur 2021;1:100012. https://doi.org/10.1016/j.lanepe.2020.100012.

[39] Thunstrom L, Ashworth M, Finnoff D, Newbold S. Hesitancy towards a COVID-19 vaccine and prospects for herd immunity. SSRN Electron J 2020. https://doi.org/10.2139/ssrn.3593098.

[40] Megget K. Even covid-19 can’t kill the anti-vaccination movement. BMJ 2020;369:m2184. https://doi.org/10.1136/bmj.m2184.

[41] Kasozi KI, Laudisoit A, Osuwat LO, Batiha GES, Al Omairi NE, Aigbogun E, et al. A descriptive-multivariate analysis of community knowledge, confidence, and trust in covid-19 clinical trials among healthcare workers in Uganda. Vaccines 2021. https://doi.org/10.3390/vaccines9030253.

[42] Freeman D, Loe BS, Chadwick A, Vaccari C, Waite F, Rosebrock L, et al. COVID-19 vaccine hesitancy in the UK: The Oxford coronavirus explanations, attitudes, and narratives survey (Oceans) II. Psychol Med 2021:1–15. https://doi.org/10.1017/S0033291720005188.

[43] Anaki D, Sergay J. Predicting health behavior in response to the coronavirus disease (COVID-19): Worldwide survey results from early March 2020. PLoS One 2021;16. https://doi.org/10.1371/journal.pone.0244534.

[44] Karlsson LC, Soveri A, Lewandowsky S, Karlsson L, Karlsson H, Nolvi S, et al. Fearing the disease or the vaccine: The case of COVID-19. Pers Individ Dif 2021;172:110590. https://doi.org/10.1016/j.paid.2020.110590.

[45] Chen Y, Zhou R, Chen B, Chen H, Li Y, Chen Z, et al. Knowledge, perceived beliefs, and preventive behaviors related to covid-19 among Chinese older adults: Cross-sectional web-based survey. J Med Internet Res 2020;22. https://doi.org/10.2196/23729.

[46] Fisk RJ. Barriers to vaccination for coronavirus disease 2019 (COVID-19) control: experience from the United States. Glob Heal J 2021. https://doi.org/10.1016/j.glohj.2021.02.005.

[47] Saied SM, Saied EM, Kabbash IA, Abdo Saef. Vaccine hesitancy: Beliefs and barriers associated with COVID-19 vaccination among Egyptian medical students. J Med Virol 2021:1–12. https://doi.org/10.1002/jmv.26910.

[48] Wong MCS, Wong ELY, Huang J, Cheung AWL, Law K, Chong MKC, et al. Acceptance of the COVID-19 vaccine based on the health belief model: A population-based survey in Hong Kong. Vaccine 2021. https://doi.org/10.1016/j.vaccine.2020.12.083.

[49] Wong LP, Alias H, Wong PF, Lee HY, AbuBakar S. The use of the health belief model to assess predictors of intent to receive the COVID-19 vaccine and willingness to pay. Hum Vaccines Immunother 2020;16:2204–14. https://doi.org/10.1080/21645515.2020.1790279.

